# Doxycycline prophylaxis for sexually transmitted infection prevention in Vietnam: Awareness, attitudes, and willingness to use among men who have sex with men using HIV-PrEP

**DOI:** 10.1101/2024.12.02.24318296

**Authors:** HTM Bui, PC Adamson, JD Klausner, GM Le, PM Gorbach

## Abstract

**Objectives:** Doxycycline prophylaxis shows promise for sexually transmitted infection (STI) prevention, but data primarily come from high-income countries. This study assessed awareness, willingness and factors associated with willingness to use doxycycline for STI prevention among men who have sex with men (MSM) using HIV-PrEP in Vietnam.

**Methods:** Between January 25 and February 4, 2024, a cross-sectional study recruited males aged ≥18 years who reported sex with men in the past 12 months from 11 PrEP clinics in Hanoi and Ho Chi Minh City. Self -administered surveys were conducted, and multivariable logistic regression was applied to identify factors associated with willingness to use doxyPEP/PrEP.

**Results:** Among 350 participants (45.7% Hanoi, 54.3% Ho Chi Minh City), the median age was 25 (interquartile range [IQR]: 21-30), and 10.6% self-reported an STI diagnosis in the past 12 months. In the previous 6 months, the median number of sex partners was 2 (IQR: 1–4), 53.1% reported condomless anal sex. Awareness of doxycycline was low (20.2%; 65/322), however, 75.4% (264/350) expressed willingness to use it, with 63.6% (168/264) preferring doxyPrEP. Participants who disclosed HIV-PrEP use to all sex partners (adjusted odds ratio [aOR]: 4.17; 95% confidence interval [95%CI] 1.84, 9.46), and those with higher perceived STI risk (aOR: 1.12; 95% CI 1.03, 1.22) were more likely to report willingness to use doxycycline prophylaxis. Concerns about daily medication (aOR: 0.43; 95% CI 0.24, 0.81) and fear of judgement from peers (aOR: 0.41; 95% CI: 0.21, 0.81) were associated with lower willingness.

**Conclusions:** Knowledge of doxyPEP/PrEP among MSM on HIV-PrEP in Vietnam was low. However, most expressed willingness to use doxycycline prophylaxis, with two-thirds preferring doxyPrEP. Findings highlight the need to disseminate information on doxyPEP/PrEP for STI prevention strategy, evaluating the usage and comparative effectiveness of doxyPEP and doxyPrEP to guide implementation efforts in Vietnam.

**1. What is already known on this topic:** Men who have sex with men (MSM) experience a disproportionate burden of HIV and bacterial sexually transmitted infections (STIs) worldwide, including in Vietnam. Despite the successful expansion of HIV Pre-Exposure Prophylaxis (PrEP) programs in Vietnam, bacterial STI rates remain persistently high among PrEP users, highlighting gaps in prevention efforts. Evidence from high-income countries (HICs) suggests that doxycycline prophylaxis can significantly reduce the incidence of bacterial STIs such as syphilis and chlamydia. However, little is known about the awareness, acceptability, and feasibility of doxycycline prophylaxis in Vietnam and other low-and middle-income countries (LMICs).

**2. What this study adds:** This study is the first to assess awareness, attitudes, and willingness to use doxycycline prophylaxis for STI prevention among MSM in Vietnam, a LMIC setting. Despite low prior awareness, the study found high willingness (75.4%) to use doxycycline, with most participants preferring daily doxyPrEP over doxyPEP. It also identified critical facilitators, such as perceived high STI risk and disclosure of HIV-PrEP use to sex partners, as well as barriers, including stigma, cost, and concerns about daily medication. These findings provide insights into the feasibility of introducing doxycycline prophylaxis in Vietnam, distinguishing it from contexts in HICs where different preferences and barriers may exist.

**3. How this study might affect research, practice, or policy:** This study provides a foundation for further research on doxycycline prophylaxis in LMICs, emphasizing the need to evaluate its real-world effectiveness and implementation in settings with high AMR prevalence. Policymakers can use these findings to develop tailored guidelines and subsidies for doxycycline prophylaxis in Vietnam, ensuring affordability and accessibility.

## Background

Men who have sex with men (MSM) globally face a disproportionate burden of HIV and sexually transmitted infections (STIs) including syphilis, chlamydia and gonorrhoea.^1^ In Vietnam, MSM have high prevalences of HIV (10.9%), syphilis (18.9%), chlamydia (23.9%), and gonorrhea (13.9%) infections.^2^ In 2018, the national HIV pre-exposure prophylaxis (PrEP) program first became available in Vietnam. While PrEP is highly effective for HIV prevention, bacterial STIs among PrEP participants remain high.^3^ A study conducted among MSM receiving HIV-PrEP at a sexual health clinic in Hanoi in 2022 revealed that 29.3% of participants were infected with *C. trachomatis* or *N.gonorrhoeae*.^4^ These findings underscore the urgent need for STI prevention interventions in this population.

Clinical trials have consistently demonstrated the effectiveness of doxycycline prophylaxis in preventing syphilis, and chlamydia among MSM.^5–7^ Doxycycline prophylaxis for STI prevention can be administered as post-exposure prophylaxis (referred to as doxyPEP) taken within 72 hours of condomless sex, or as daily prophylaxis (referred to as doxyPrEP). Doxycycline, a commonly used second-generation tetracycline, is associated with good tolerability and a low risk of serious side effects.^8^ Data from clinical trials^5, 6, 9^ and a systematic review of 67 studies^10^ confirm the safety of doxycycline event with longer-term use, defined as greater than eight weeks, and reported minimal side-effects.

Given the effectiveness of doxycycline for STI prevention, health departments in Europe, the US and Australia^11–13^ have published guidelines for health care workers to consider or recommend its use for populations at high risk for STIs, including MSM. In Vietnam, doxycycline is widely available for treatment of chlamydia, syphilis and other infections; however, no guidelines exist for doxyPEP/PrEP.

Most studies on willingness to use doxycycline and potential concerns have been conducted in high-income countries, showing high acceptability and feasibility.^14–16^ Data from low-and middle-income countries (LMICs) are scarce. A clinical trial of doxycycline prophylaxis for STI prevention among cisgender women in Kenya, the only done in a LMIC, found doxycycline ineffective for STI prevention, likely due to low adherence.^9^ This highlights the need for research in diverse settings, where antibiotic use behaviors and acceptability may differ. Understanding community willingness and concerns is crucial for predicting uptake and informing implementation strategies. We conducted a survey among MSM using HIV-PrEP in clinics in two cities in Vietnam to evaluate their knowledge, interest, and attitudes, as well as factors associated with willingness to take doxycycline prophylaxis for STI prevention.

## Methods

### Design

This cross-sectional study included 350 MSM attending 11 clinics for HIV-PrEP follow-up and uptake. Since 2019, the national HIV-PrEP program has been provided free of charge at 210 clinics across 29 of 63 provinces in the country. Over 80,000 patients have initiated HIV-PrEP, with 80.2% of whom are MSM^17^. The sample size was calculated based on an estimated willingness to use doxycycline for STI prevention (60% - 70% in the US and Australia),^14, 15^ a precision of 10%, and a population size of MSM 64,000 MSM ever on HIV-PrEP in Vietnam.^17^

### Setting

The study was conducted at 6 clinics in Hanoi, the capital city in the North, and 5 clinics in Ho Chi Minh City, the most populous Southern city. Hanoi and Ho Chi Minh City have the highest number of HIV-PrEP clients in Vietnam. Both public and private clinics with 80% PrEP male clients reported having sex with other men were selected. The survey was conducted over 11 days, from January 25 to February 4, 2024.

### Participants

Eligible participants were male aged 18 or older, who had sex with at least one man in the past 12 months, had taken oral HIV-PrEP for at least 30 days and attended a clinic for a HIV-PrEP follow-up visit.

Study recruitment used posters and flyers at the 11clinics. Clinic staff pre-screened and introduced the study to potential participants. Those interested in participating were referred to a research assistant in a private room for eligibility confirmation, study explanation, and written informed consent.

The study was reviewed and approved by IRB at Hanoi Medical University (1102/GCN-HDDDNCYSH-DHYHN). All participants provided informed consent prior to participating in the study.

### Data collection and measurement

A tablet-based self-administerd questionnaire was used to collect demographics, sexual and substance use behaviors, HIV-PrEP use, knowledge and perception of STIs, and of doxycycline use for STI prevention. The questionnaire was piloted with clients from 11 clinic for revising and revised before implementation. Participants were instructed on using the tablet for self-administering the questionnaire. A research assistant remained available outside the room for questions that arose. Upon completion of the survey, which required appoximately 20 to 30 minutes, participants were compensated with 150,000 VND (equivalent to 7 USD) for their time.

The primary outcome was willingness to use doxycycline for STI prevention, assessed by asking ‘If your physician doxycycline for STI prevention, would you take it?’. Responses were on a 5-point Likert scale from “Definitely no” to “Definitely yes,”. Participants who responded “Definitely Yes” and “Yes’ were classified as “willing” to take doxyPEP/PrEP; while others were classified as “not willing.”

We reported socio-demographic data (age, education, employment, monthly income) and assessed social engagement with gay men using the validated Australian *Gay Social Engagement* scale.^18^ Score of the scale ranged between 0 and 9, and was calculated by summing numerically coded responses to two survey questions: the number of gay friends, and the amount of free time spent with gay men. Sexual perception and practices included sexual orientation, preferred sexual positioning, number of male sex partners and condomless anal sex in the previous 6 months. Participants answered (yes/no) to questions about sex with female and transgender partners, group sex, sex while using substances such as party drugs, popper/Viagra, amphetamines-type stimulants (ATS) to enhance sexual performance, and sex with partners met via websites or apps in the previous 6 months. Those who reported having sex while using party drugs, using popper/Viagra or ATS to enhance sexual performance was categorized as engaging in ‘chemsex’. Substance use in the previous 6 months was reported.

We collected experience related to HIV-PrEP, including time since initiation, regimens used in the previous 12 months (consistent daily use, periodic daily use (during period of risk^19^), event-driven, and a mix of regimens), disclosure of HIV-PrEP use to male sex partner, and perception of less condom use among MSM using HIV-PrEP.

Participants’ knowledge of STIs was assessed using three questions for each disease: 1-Is syphilis/ chlamydia/ gonorrhea an STI (yes/ no/ don’t know); 2-What causes the infection (bacteria/virus/don’t know); 3-Can it be cured (yes/ no/don’t know). Each correct answer scored 1 point; incorrect or ‘don’t know’ responses scored 0, yielding a total STI knowledge score from 9 questions. Participants self-rated their STI risk on a 0-10 scale, with a higher score indicating higher perceived risk. They also rated the STI risk for MSM using HIV-PrEP on a 5-level scale. Participants reported lifetime STI testing, STI diagnoses in the previous 12 months, and any anal or genital STI-related symptoms.

Participants received brief information on doxycycline, including its prescription practice, cost, tolerability, common gastrointestinal side effects, concerns about its use, and effectiveness for STI prevention. Data were then collected on participants’ prior knowledge of doxycycline for STI prevention, willingness to use it if prescribed, preferences for PrEP/PEP and concerns about its use.

### Statistical Analysis

Data were summarized as mean and standard deviation (SD) for normally distributed continuous variables, or median and interquartile range (IQR) for abnormally distributed continuous ones, and as frequency and percentage for categorical variables. Differences between participants with and without willingness to use doxyPEP/PrEP were assessed using Chi-squared tests for categorical independent variables or Student’s T-tests for continuous independent variables. Multivariable logistic regression models were employed to identify factors associated with the willingness to use doxyPEP/PrEP. Predictor variables were screened using single-variable logistic regression, and those with p < 0.20 or considered confounders were included in multivariate analyses. Forward stepwise regression was used to add related variables to the final multivariate model. Unadjusted and adjusted odds ratios (aOR) and 95% confidence intervals were reported. Analyses were performed using STATA version 18 (Stata Corporation, College Station, TX, USA), with p-value < 0.05 considered statistically significant.

## Results

Table 1 presents demographic, behavioral characteristics, and HIV PrEP experience by willingness to use doxycycline for STI prevention. Of the 350 MSM enrolled in the study, 45.7% (160/350) were from Hanoi, and 54.3% (190/350) were from Ho Chi Minh City. The median age was 25 years [IQR: 21–30], and the median monthly income was 10 million VND [IQR: 5–15 million] (approximately 409.4 USD [IQR: 204.7–614.1]]. More than three quarters of the participants self-identified as gay (78.6%), with 48.6% reporting 2–5 male sex partners and 12.3% reporting >5 partners in the past 6 months. Condomless anal sex was reported by 51.7%, while 12.9% and 2.0% reported sex with female and transgender partners, respectively. In the prior 6 months, 15.4% engaged in group sex, 62.6% in ‘chemsex’, and 48.0% met partners via mobile apps. Regarding substance use, 74.9% reported alcohol use, 62.0% poppers, and 15.7% Viagra in the last 6 months. Among participants, 49.7% used HIV-PrEP for < 12 months, 70.0% used consistent daily regimen, 4.0% used periodic daily, 13,4% used event-driven, and 12.6% switched between regimens. In the past year, 65.4% disclosed HIV-PrEP use to all sex partners, 25.1% to some, and 9.4% to none. Overall, 62.0% agreed that “there is less condom use among MSM using HIV-PrEP”.

**Table 1:** Demographic, behavioral characteristics, and HIV PrEP experience of men who have sex with men using PrEP by willingness to use doxycycline for sexually transmitted infection prevention at 11 clinics in Hanoi and Ho Chi Minh City, Vietnam, 2024 (n = 350).

|  | Total<br>N (%) | Willingness to use doxycycline for<br>STI prevention (n=350) |  | p-value |
| --- | --- | --- | --- | --- |
|  |  | Yes (n=264)<br>N (%) | No (n=86)<br>N (%) |  |
| **SOCIAL-DEMOGRAPHIC CHARACTERISTICS** |  |  |  |  |
| **City** |  |  |  | 0.409 |
| Hanoi | 160 (45.7) | 124 (47) | 36 (41.9) |  |
| Ho Chi Minh | 190 (54.3) | 140 (53) | 50 (58.1) |  |
| **Age (median, IQR)** | 25 [21,30] | 25 [22,30] | 25 [21,30] | 0.906 |
| 18 – < 25 years old | 169 (48.3) | 127 (48.1) | 42 (48.8) |  |
| 25+ years old | 181 (51.7) | 137 (51.9) | 44 (51.2) |  |
| **Educational level** |  |  |  | 0.551 |
| High school or below | 56 (16.0) | 44 (16.7) | 12 (14) |  |
| Attending/completed college,<br>university or higher | 294 (84.0) | 220 (83.3) | 74 (86) |  |
| **Employment status** |  |  |  | 0.636 |
| Full-time employed | 226 (64.6) | 167 (63.3) | 59 (68.6) |  |
| Part-time employed | 65 (18.6) | 53 (20.1) | 12 (14) |  |
| Unemployed | 54 (15.4) | 40 (15.2) | 14 (16.3) |  |
| Not working | 5 (1.4) | 4 (1.5) | 1 (1.2) |  |
| **Monthly personal income (VND)** | 10 [6,15] | 10 [6,15] | 10 [6,15] | 0.844 |
| ≤ 7 million | 117 (33.4) | 89 (33.7) | 28 (32.6) |  |
| > 7 million | 233 (66.6) | 175 (66.3) | 58 (67.4) |  |
| **Gay community engagement score** | 5.3 ± 1.4 | 5.3 ± 1.4 | 5.1 ± 1.4 | 0.191 |
| **SEXUAL PERCEPTION AND PRACTICES** |  |  |  |  |
| **Self-identify sexual orientation** |  |  |  | 0.402 |
| Heterosexual | 10 (2.9) | 6 (2.3) | 4 (4.7) |  |
| Gay | 275 (78.6) | 212 (80.3) | 63 (73.3) |  |
| Bisexual | 63 (18) | 45 (17) | 18 (20.9) |  |
| Asexual | 2 (0.6) | 1 (0.4) | 1 (1.2) |  |
| **Reported preference sexual position** |  |  |  | 0.623 |
| Insertive | 117 (33.4) | 87 (33) | 30 (34.9) |  |
| Receptive | 87 (24.9) | 69 (26.1) | 18 (20.9) |  |
|  |  | Yes (n=264)<br>N (%) | No (n=86)<br>N (%) |  |
| **Had no preference for sexual position** | 146 (41.7) | 108 (40.9) | 38 (44.2) |  |
| **Number of male sex partners, past 6 months (median, IQR)** | 2 [1,4] | 2 [1,4] | 2 [1,3] | 0.486 |
| **1** | 137 (39.1) | 103 (39) | 34 (39.5) | 0.203 |
| **2-5** | 170 (48.6) | 124 (47) | 46 (53.5) |  |
| **>= 5** | 43 (12.3) | 37 (14) | 6 (7) |  |
| **Condomless in anal sex with male sex partners, past 6 months** | 181 (51.7) | 132 (50) | 49 (43) | 0.261 |
| **Had sex with female partners in the last 6 months** | 45 (12.9) | 30 (11.4) | 15 (17.4) | 0.144 |
| **Had sex with transgender partners in the last 6 months** | 7 (2) | 2 (0.8) | 5 (5.8) | **0.004** |
| **Group sex, past 6 month** | 54 (15.4) | 42 (15.9) | 12 (14) | 0.663 |
| **Sexualized drug use, past 12 months** | 219 (62.6) | 173 (65.5) | 46 (53.5) | **0.045** |
| **Had sex partners met via mobile apps** | 168 (48) | 127 (48.1) | 41 (47.7) | 0.945 |
| **SUBSTANCE USE PRACTICES** |  |  |  |  |
| **Substance use in the past 6 months** |  |  |  |  |
| **Alcohol** | 262 (74.9) | 203 (76.9) | 59 (68.6) | 0.124 |
| **Popper** | 217 (62) | 172 (65.2) | 45 (52.3) | **0.033** |
| **Laughing gas** | 41 (11.7) | 32 (12.1) | 9 (10.5) | 0.678 |
| **Viagra “Thai house”** | 55 (15.7) | 41 (15.5) | 14 (16.3) | 0.868 |
| **Bath salts (Mephedrone)** | 3 (0.9) | 2 (0.8) | 1 (1.2) | 0.723 |
| **Club drug (GHB)** | 10 (2.9) | 5 (1.9) | 5 (5.8) | 0.058 |
| **Amphetamine/Meth (Crystal, MDMA, ecstasy)** | 13 (3.7) | 7 (2.7) | 6 (7) | 0.069 |
| **HIV-PrEP USE EXPERIENCE AND PERCEPTION** |  |  |  |  |
| **Time since PrEP initiation** |  |  |  | 0.186 |
| **≤ 12 months** | 174 (49.7) | 135 (51.1) | 39 (45.3) |  |
| **>1 year – 3 years** | 119 (34) | 83 (31.4) | 36 (41.9) |  |
| **>3 years** | 57 (16.3) | 46 (17.4) | 11 (12.8) |  |
| **HIV-PrEP regimen in the last 12 months** |  |  |  | 0.123 |
| **Consistent daily regimen** | 245 (70) | 187 (70.8) | 58 (67.4) |  |
| **Periodic daily regimen** | 14 (4) | 8 (3) | 6 (7) |  |
|  |  | Yes (n=264)<br>N (%) | No (n=86)<br>N (%) |  |
| **Event-driven (ED) regimen** | 47 (13.4) | 32 (12.1) | 15 (17.4) |  |
| **Mixed regimen** | 44 (12.6) | 37 (14) | 7 (8.1) |  |
| **Disclosed HIV-PrEP use to male sex**<br>**partners, past 12 months** |  |  |  | **<0.001** |
| **No** | 33 (9.4) | 18 (6.8) | 15 (17.4) |  |
| **Disclosed to some male partners** | 88 (25.1) | 58 (22) | 30 (34.9) |  |
| **Disclosed to all male partners** | 229 (65.4) | 188 (71.2) | 41 (47.7) |  |
| **Perception there is less condom use**<br>**among MSM using HIV-PrEP** |  |  |  | 0.271 |
| **Agree** | 217 (62) | 170 (64.4) | 47 (54.7) |  |
| **Neither** | 99 (28.3) | 70 (26.5) | 29 (33.7) |  |
| **Disagree** | 34 (9.7) | 24 (9.1) | 10 (11.6) |  |

Of all participants, 264 (75.4%) were willing to take doxycycline for STI prevention. Those willing to use doxycycline were less likely to report having transgender sex partners (0.8% vs. 5.8%, p=0.004) but more likely to engage in ‘chemsex’ (65.5% vs. 53.5%, p=0.045) and use poppers (65.2% vs. 52.3%, p=0.033) in the prior 6 months, compared to those with low willingness to use (Table 1). Among those willing to use doxycycline, 6.8% did not disclose their HIV-PrEP use to sex partners, compared to 17.4% among those not willing to use doxycycline (p<0.001).

Knowledge, perception, experience with STIs and prevention are shown in Table 2. The median self-perceived risk score for STI was 6.0/10.0 overall, with those willing to use doxycycline reporting higher perceived STI risk (6.0 vs. 4.0, p=0.009) compared to those without willingness. Syphilis testing was reported by 80.6%, followed by 59.7% for gonorrhea, and 30.0% for chlamydia. In the previous 12 months, 0.6% self-reported being diagnosed with syphilis, 2.6% with chlamydia, and 8.3% gonorrhea. There was no difference in willingness to use doxycycline among those who had ever tested or been diagnosed with STIs in the prior 12 months or had anal/urethral STI symptoms in the prior 6 months.

**Table 2:** Knowledge, perception, experience with STIs and STI prevention among men who have sex with men using HIV PrEP by willingness to use doxycycline for STI prevention at 11 clinics in Hanoi and Ho Chi Minh City, Vietnam, 2024 (n = 350).

|  | Total<br>N (%) | Willingness to uptake Doxycycline<br>for STI prevention (n=350) |  | p-value |
| --- | --- | --- | --- | --- |
|  |  | Yes (n=264)<br>N (%) | No (n=86)<br>N (%) |  |
| STI knowledge score | 7 [6,8] | 7 [6,8] | 7 [5,8] | 0.673 |
| Self-perceived risk score for STIs | 6 [3,9] | 6 [3,9] | 4 [2,8] | **0.009** |
| Perception that MSM have high risk for STIs | 207 (59.1) | 156 (59.1) | 51 (59.3) | 0.972 |
| Ever tested for |  |  |  |  |
| Syphilis | 282 (80.6) | 214 (81.1) | 68 (79.1) | 0.614 |
| Gonorrhea | 209 (59.7) | 158 (59.8) | 51 (59.3) | 0.519 |
| Chlamydia | 105 (30) | 82 (31.1) | 23 (26.7) | 0.690 |
| Any of these STIs | 292 (83.4) | 222 (84.1) | 70 (81.4) | 0.422 |
| Diagnosed with STIs in prior 12 months |  |  |  |  |
| Syphilis | 2 (0.6) | 1 (0.4) | 1 (1.2) | 0.402 |
| Gonorrhea | 9 (2.6) | 6 (2.3) | 3 (3.5) | 0.536 |
| Chlamydia | 29 (8.3) | 19 (7.2) | 10 (11.6) | 0.195 |
| Any of these STIs | 37 (10.6) | 25 (9.5) | 12 (14.0) | 0.240 |
| Reported anal/genital STI symptoms,<br>prior 6 months | 79 (22.6) | 61 (23.1) | 18 (20.9) | 0.675 |
| Heard of doxycycline prophylaxis for STI<br>prevention (n=322) | 65 (20.2) | 42 (17.4) | 23 (28.4) | **0.033** |
| Doxycycline regimen preference for STI<br>prevention |  |  |  | 0.200 |
| PrEP | 272 (77.7) | 207 (78.4) | 65 (75.6) |  |
| PEP | 77 (22) | 57 (21.6) | 20 (23.3) |  |
| Neither | 1 (0.3) | 0 (0) | 1 (1.2) |  |
| Concerns about doxycycline use for STI<br>prevention |  |  |  |  |
| Cost of the medicine | 122 (34.9) | 97 (36.7) | 25 (29.1) | 0.195 |
| Must take the medicine daily | 76 (21.7) | 47 (17.8) | 29 (33.7) | **0.002** |
| Must take the medicine within 72 hours<br>after having sex | 73 (20.9) | 51 (19.3) | 22 (25.6) | 0.214 |
|  |  | Yes (n=264)<br>N (%) | No (n=86)<br>N (%) |  |
| **Complications of the medicine**<br>**(antimicrobial resistance, symptoms...)** | 213 (60.9) | 157 (59.5) | 56 (65.1) | 0.351 |
| **Judgement from peers** | 52 (14.9) | 29 (11) | 23 (26.7) | <0.001 |

There were 20.2% (65/322) of participants who reported ever hearing of doxycycline for STI prevention, and only one had previously used it for this purpose. The most common concerns regarding doxycycline uptake were potential side effects and antimicrobial resistance (60.9%), followed by cost (34.9%). Among those willing to use doxycycline, fewer had heard of it for STI prevention (17.4% vs. 28.4%, p=0.033), reported worry about daily usage (17.8% vs. 33.7%, p=0.002), and concerned about judgement from peers (11.0% vs. 26.7%, p<0.001) (Table 2).

Figure 1 presents awareness and willingness to use doxycycline prophylaxis stratified by self-reported STI diagnosis. There were no significant differences in awareness or willingness to use doxycycline between those with and without a recent STI diagnosis.

**Figure 1:**
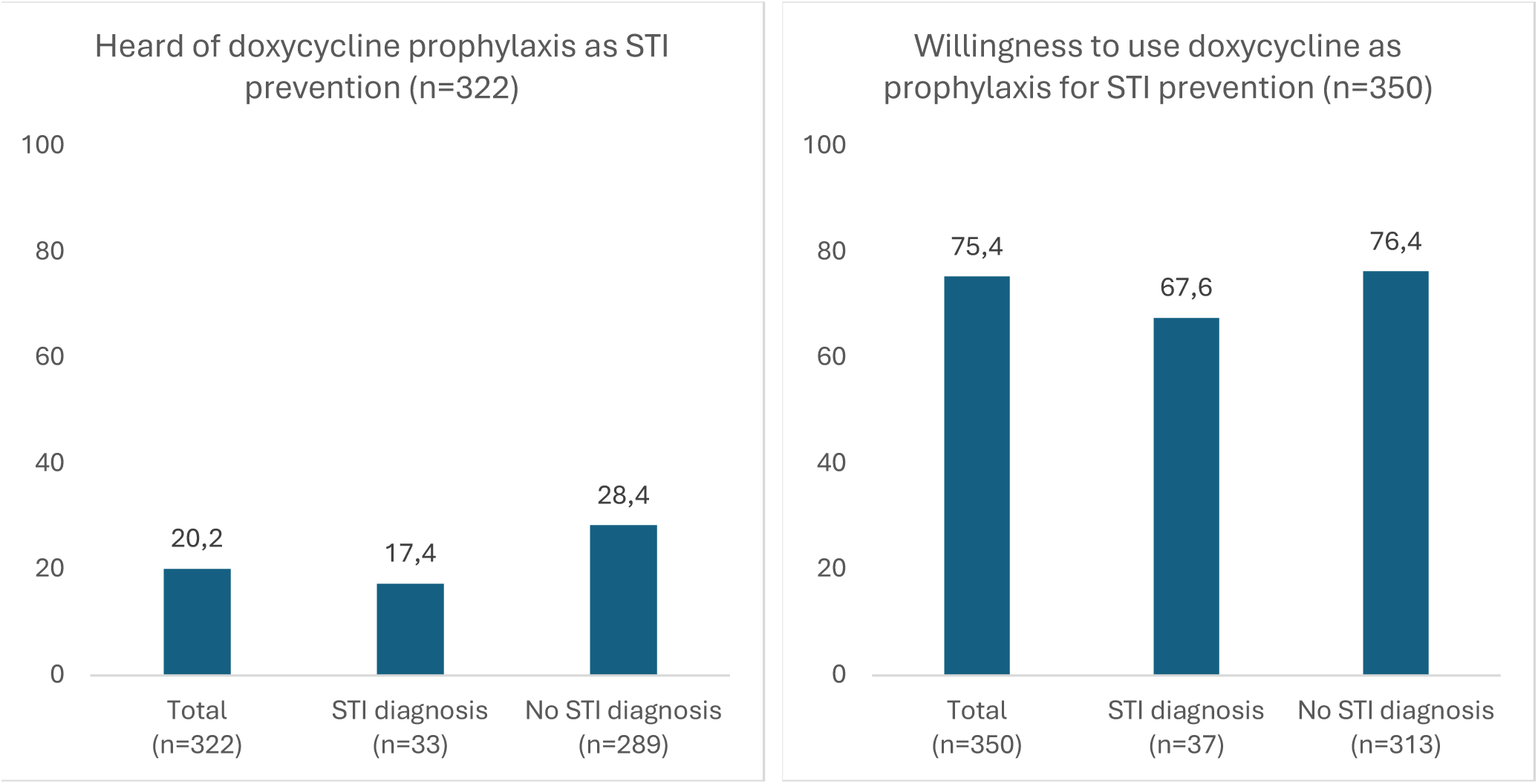
Awareness and willingness to use doxycycline among men who have sex with men using HIV PrEP by self-reported STI diagnosis in the previous 12 months.

In multivariate analysis, controlling for age, education, and study site, factors independently associated with willingness to use doxycycline for STI prevention included HIV-PrEP disclosure to all male sex partners (aOR=4.40, 95%CI 1.94-9.97) and higher self-perceived STI risk (aOR=1.14, 95%CI 1.05-1.24). Concern about daily medication use (aOR=0.43, 95%CI 0.23-0.79) and judgement from peers (aOR=0.40, 95%CI 0.20-0.79) were associated with lower willingness. Use of poppers in the prior 6 months was associated with willingness to use doxycycline in the univariate model, but not in the multivariate model. (Table 3)

**Table 3.** Logistic regression models of factors associated with willingness to use doxycycline for STI prevention among men who have sex using PrEP at 11 clinics in Hanoi and Ho Chi Minh City, Vietnam, 2024 (n = 350).

|  | Willingness to use /<br>row N |  | Unadjusted Rate<br>Ratio [95%CI] | Adjusted Rate<br>Ratio [95%CI] |
| --- | --- | --- | --- | --- |
|  | n/N | % |  |  |
| **Overall** | 264/350 | 75.4 |  |  |
| **City** |  |  |  |  |
| **Hanoi** | 124/160 | 77.5 | 1.00 | 1.00 |
| **Ho Chi Minh** | 140/190 | 73.7 | 0.81 [0.50,1.33] | 0.85 [0.49,1.48] |
| **Age** |  |  |  |  |
| **18 – < 25 years old** | 127/169 | 75.1 | 1.0 | 1.0 |
| **25+ years old** | 137/181 | 75.7 | 1.03 [0.63,1.67] | 0.98 [0.57,1.69] |
| **Educational level** |  |  |  |  |
| **High school or below** | 44/56 | 78.6 | 1.00 | 1.00 |
| **Attending/completed university**<br>**or higher** | 220/294 | 74.8 | 0.81 [0.41,1.62] | 0.60 [0.28,1.30] |
| **Popper use, past 6 months** | 172/217 | 79.3 | **1.70 [1.04,2.79]** | 1.52 [0.96,2.86] |
| **Disclosed PrEP use to male sex**<br>**partners, past 12 months** |  |  |  |  |
| **No** | 18/33 | 54.5 |  | 1.00 |
| **Disclosed to some male**<br>**partners** | 58/88 | 65.9 | 1.61 [0.71,3.64] | 1.60 [0.67,3.81] |
| **Disclosed to all male partners** | 188/229 | 82.1 | 3.82 [1.78,8.20] | **4.17 [1.84,9.46]** |
| **Self-perceived risk score for STIs** |  |  | 1.11 [1.03,1.19] | **1.12 [1.03,1.22]** |
| **Ever heard of doxycycline use for**<br>**STI prevention (n=322)** | 42/65 | 64.6 | 0.98 [0.71,1.36] |  |
| **Worried about doxycycline use**<br>**for STI prevention** |  |  |  |  |
| **Must take the medicine daily** | 47/76 | 61.8 | 0.43 [0.25,0.74] | **0.43 [0.24,0.81]** |
| **Judgement from peers** | 29/52 | 55.8 | 0.34 [0.18,0.62] | **0.41 [0.21,0.81]** |

**Bolded values** – denote those associated with p-values <0.05.

## Discussion

We conducted a survey with MSM attending HIV-PrEP clinics in the two largest cities in Vietnam to assess their awareness, attitudes, and willingness to use doxycycline for STI prevention. While prior knowledge of doxycycline prophylaxis was low, willingness to use it was high among MSM on HIV-PrEP, a targeted population for doxycycline use in other countries’ guidelines.^11–13^ Notably, doxyPrEP was preferable to doxyPEP, which was surprising and provides insights into preferences of this population. Given the limited data on doxycycline prophylaxis from LMICs, our findings are valuable for planning its implementation in LMICs settings.

The high acceptability of doxycycline prophylaxis among MSM using HIV-PrEP aligns with previous studies, which reported willingness to use it ranging from 52.7% to 90.6%, suggesting the uptake will be likely high in Vietnam.^14–16^ Studies have shown a high prevalence of bacterial STIs among MSM, both in the community ^20^ and those on HIV PrEP,^21^ highlighting the urgent need doxycycline access. However, we found low self-reported recent STI diagnoses, and this was not associated with willingness to use, which is contrary to some reports.^14–16^ This could be due to self-report bias, or limited STI testing access. It also may reflect our study sample that was MSM on HIV PrEP who may be already engaged in prevention programs and so have more access to STI testing and treatment. Despite national PrEP guidelines recommending quarterly STI testing,^22^ national HIV-PrEP program data showed coverage of STI testing was suboptimal.^23^ Survey data estimate that up to 10% of MSM in the UK, Netherlands, and Australia used antibiotic STI prophylaxis without appropriate guidance between 2018 and 2020, emphasizing the need for clear, accessible information in countries like Vietnam, where no formal guidelines exist.^24–26^ Given the low prior knowledge observed, it is crucial to disseminate information about doxycycline prophylaxis to MSM using HIV-PrEP, their providers, and other populations who may benefit. Innovative community-based strategies, such as crowdsourcing and co-creation^27^, which have proven effective in other public health initiative,^27^ could support successful implementation.

Our study highlights potential barriers to doxycycline prophylaxis implementation for STI prevention in Vietnam, with primary concerns being side effects and AMR. While concerns about doxycycline’s complications are understandable, evidence shows it is generally safe with minimal adverse effects during long-term use^10^. A recent review found no doxycycline resistance for syphilis and chlamydia, but further research is needed to assess AMR risk for *Neisseria gonorrhoeae* and commensal *Neisseria* species.^28^ Randomized controlled trials (RCTs) have shown significant reductions in syphilis and chlamydia, though data on gonorrhea have been mixed due to tetracycline resistance in *N. gonorrhoeae*.^6, 7^ In Vietnam, where tetracycline resistance in *N. gonorrhoeae* is high (80-90%), the effectiveness of doxycycline for gonorrhea remains uncertain.^29^ As AMR in *N. gonorrhoeae* is a growing concern in Vietnam and the region, the potential risks of AMR in *N. gonorrhoeae*, as well as other pathogens, ^29^ must be weighed against the benefits of reducing STI incidence with doxyPEP/PrEP. Cost was another concern in our study, but doxycycline is available as a generic, and government subsidies could help enhance uptake in other LMICs.

Our findings that participants with higher perceived STI risk scores were more willing to use the medication aligns with previous studies among MSM in the US.^14^ However, the reduced willingness among those concerned about judgment from peers represents a novel finding. This association, observed in our population of MSM using HIV-PrEP in Vietnam, may also be relevant in other contexts with high stigma towards MSM and STIs.^30^ Future doxyPEP/PrEP implementation of in countries with significant stigma among MSM should include interventions to reduce perceived stigma and enhance self-esteem among MSM.

Our study should be interpreted considering the following limitations. The survey was conducted over 11 days to recruit MSM who were visiting clinics for on-site HIV-PrEP refills and could not reach those who missed appointment or were lost-to-follow-up, which may limit the representativeness of MSM using HIV-PrEP in Vietnam. MSM not using HIV-PrEP were also not included, potentially missing individuals at risk for HIV and STIs. Self-reported responses may have introduced social desirability and recall biases, which might impact the null and limit the observed effect size. Additionally, the survey presented a hypothetical intervention and used a structured questionnaire without open-ended options, potentially limiting insight into subjective concerns. Despite these limitations, this is the first study in Vietnam to examine awareness, and willingness to use doxycycline for STI prevention among MSM using HIV-PrEP. In conclusion, most participants expressed willingness to use doxycycline prophylaxis for STI prevention, with nearly two thirds preferring daily PrEP, despite low prior knowledge about doxycycline among MSM on HIV-PrEP in Vietnam. These findings underscore the need for guidance on implementing doxyPEP/PrEP in Vietnam and emphasize broader dissemination of information about doxycycline as an STI prevention strategy. Future efforts should explore offering both PrEP/PEP options, targeting population most likely to benefit, and addressing factors influencing willingness to use doxycycline prophylaxis. Findings from this study can inform policymakers about the need for STI prevention strategies for MSM in Vietnam and might be important for other LMICs.

## Data Availability

All data produced in the present study are available upon reasonable request to the authors

## Acknowledgements

We are grateful to the study participants and clinic staff at 11 HIV-PrEP clinics in Vietnam for their support. We extend our special thanks to Mr. Bao An, Director of the Center for Applied Research on Men and Community Health (CARMAH) for his critical role in facilitating the survey implementation at five clinics in Ho Chi Minh City. We also express our appreciation to our colleagues -Mr Bui Trung Thanh, Dr. Pham Quang Loc, Ms Nguyen Thi Phuong Linh, Mr. Nguyen Cong Thanh and Mr. Dau Sy Nguyen - at the Center for Training and Research on Substance Abuse – HIV (CREATA-H) and the Sexual Health Promotion (SHP) clinic at Hanoi Medical University (HMU), for their valuable consultation and support in survey implemenation and data management. Last, we are thankful to students at the Hanoi Medical University, and the University of Medicine and Pharmacy at Ho Chi Minh City for their assistance with data collection.

## Contributors

Study conceptualization and methodology: HTMB, PCA, KJD, GML, PMG. Data curation and analysis: HTMB. Writing – original draft preparation, editing and finalization; HTMB. Writing: review and revision with important intellectual contributions: PCA, KJD, PMG. All authors reviewed and approved the manuscript.

## Patient consent for publication

Not applicable.

## Data availability statement

Data are available upon reasonable request. Deidentified individual participant data and other supporting documents will be made available upon reasonable requests made to the corresponding author.

